# The Diagnostic Accuracy of Subjective Dyspnea in Detecting Hypoxemia Among Outpatients with COVID-19

**DOI:** 10.1101/2020.08.10.20172262

**Authors:** Linor Berezin, Alice Zhabokritsky, Nisha Andany, Adrienne K. Chan, Andrea Gershon, Philip W. Lam, Jerome A. Leis, Scott MacPhee, Samira Mubareka, Andrew E. Simor, Nick Daneman

**Affiliations:** Faculty of Medicine, University of Toronto, Toronto, Canada; Division of Infectious Diseases, Sunnybrook Health Sciences Centre, University of Toronto; Division of Respirology, Sunnybrook Health Sciences Centre, University of Toronto; Department of Nursing, Sunnybrook Health Sciences Centre, University of Toronto; Department of Laboratory Medicine and Pathology, University of Toronto, Toronto, Canada

**Keywords:** COVID-19, Dyspnea, hypoxemia, oxygen saturation, monitoring, virtual care

## Abstract

**Objectives:** The majority of patients with mild-to-moderate COVID-19 can be managed using virtual care. Dyspnea is challenging to assess remotely, and the accuracy of subjective dyspnea measures in capturing hypoxemia have not been formally evaluated for COVID-19. We explored the accuracy of subjective dyspnea in diagnosing hypoxemia in COVID-19 patients.

**Methods:** This is a retrospective cohort study of consecutive outpatients with COVID-19 who met criteria for home oxygen saturation monitoring at a university-affiliated acute care hospital in Toronto, Canada from April 3, 2020 to June 8, 2020. Hypoxemia was defined by oxygen saturation <95%. Dyspnea measures were treated as diagnostic tests, and we determined their sensitivity (SN), specificity (SP), negative/positive predictive value (NPV/PPV), and positive/negative likelihood ratios (+LR/-LR) for detecting hypoxemia.

**Results:** During the study period 64/298 (21.5%) of patients met criteria for home oxygen saturation monitoring, and of these 14/64 (21.9%) were diagnosed with hypoxemia. The presence/absence of dyspnea had limited accuracy for diagnosing hypoxemia, with SN 57% (95% CI 30-81%), SP 78% (63%-88%), NPV 86% (72%-94%), PPV 42% (21%-66%), +LR 2.55 (1.3-5.1), -LR 0.55 (0.3-1.0). An mMRC dyspnea score >1 (SP 97%, 95%CI 82%-100%), Roth Maximal Count <12 (SP 100%, 95%CI 75-100%), and Roth Counting time < 8 seconds (SP 93%, 95%CI 66%-100%) had high SP that could be used to rule in hypoxemia, but displayed low SN (≤50%).

**Conclusions:** Subjective dyspnea measures have inadequate accuracy for ruling out hypoxemia in high-risk patients with COVID-19. Safe home management of patients with COVID-19 should incorporate home oxygenation saturation monitoring.

## Introduction

As of August 7, 2020, there have been more than 18 million laboratory-confirmed novel coronavirus disease 2019 (COVID-19) cases and 700,000 deaths documented globally.^1^ The spectrum of disease of COVID-19 ranges from asymptomatic or mild symptoms, to severe respiratory failure and death.^2^ Approximately 20% of patients with COVID-19 experience dyspnea, which is more commonly associated with severe disease.^3^ Fatal cases of COVID-19 have higher rates of dyspnea, lower blood oxygen saturation (SpO2), and greater rates of complications such as acute respiratory distress syndrome.^4,5^

In an effort to reduce avoidable hospitalizations, health care contacts, and transmission, most patients with COVID-19 can be managed in the community using virtual healthcare platforms, and transferred to hospital only if they develop progressive respiratory disease.6 Subjective dyspnea can be assessed remotely using patient interview, and augmented by surrogate measures such as the Roth Score and modified Medical Research Council (mMRC) Dyspnea Scale. However, the accuracy of these measures has not been formally evaluated in the context of COVID-19.6,7 Of great concern is the risk of false reassurance if patients develop hypoxemia without subjective sensation of dyspnea. “Silent hypoxemia”, or low SpO2 in the absence of dyspnea, has been reported in the setting of COVID-19 and clinicians have speculated that it may be associated with increased out-of-hospital mortality;8 case reports have described patients presenting to hospital with rapid deterioration and respiratory failure without signs of respiratory distress.^9-11^

Previous studies of the utility of dyspnea measurement in diagnosing hypoxemia in other respiratory conditions such as chronic obstructive pulmonary disease (COPD), congestive heart failure and lung cancer and have yielded conflicting results;^12-15^ This association has not been studied during the COVID-19 pandemic. Therefore, we sought to determine the diagnostic accuracy of subjective dyspnea measures in diagnosing hypoxemia among a cohort of outpatients with COVID-19.

## Methods

### Study Participants

All consecutive patients with laboratory-confirmed COVID-19 followed as outpatients by the Sunnybrook Health Sciences Centre COVID-19 Expansion to Outpatients (COVIDEO) virtual care service from April 3, 2020 to June 8, 2020 were included in this cohort study. The patients were diagnosed based on a positive mid-turbinate or nasopharyngeal swab for COVID-19 RNA detected by real-time polymerase chain reaction. COVIDEO is a virtual care model for monitoring of outpatients with COVID-19 at Sunnybrook Health Sciences Centre, and is the basis of similar programs at other hospitals.^16^ Patients were contacted by an infectious diseases physician for assessment and monitoring either by telephone or through the Ontario Telemedicine Network virtual platform.

A portable pulse oximeter was delivered to the homes of high-risk patients as defined by age > 60 years, pregnancy, extensive comorbidities, or presence of cardio-respiratory symptoms, such as chest pain or dyspnea. Patients were instructed to record their SpO2 measurements twice daily throughout their illness. This study was approved by the institutional review board at Sunnybrook Health Sciences as minimal risk research, using data collected for routine clinical practice, and the requirement for informed consent was waived.

### Data Collection

The demographic characteristics, clinical data, measures of subjective dyspnea (presence of shortness of breath, mMRC dyspnea scale score, Roth Score), physical exam findings. and SpO2 readings for study participants were collected from electronic medical records by one investigator (either A.Z or S.Ma.). The subjective dyspnea measures obtained from the patient’s first visit with a pulse oximeter were used for analysis.

### Predictor Variables

The primary predictor of interest was patient-reported presence of dyspnea. Secondary predictor variables were patient-reported breathing faster at rest, breathing harder than normal, feeling more breathless today than yesterday, as well as dyspnea as measured by the mMRC Dyspnea Scale and the Roth Score.

The mMRC Dyspnea Scale has been studied extensively in a variety of respiratory conditions.^17^ It is comprised of five categories that describe the degree of activity limitation due to worsening breathlessness. The participant assigns themselves a score ranging from 0 to 4 based on their perception of which activities result in dyspnea, with higher scores indicating a greater impairment in their ability to perform daily activities.

The Roth Score is a tool for quantifying the severity of dyspnea, in which the patient is asked to count audibly to 30 in their native language, and the maximal count and counting time are recorded. A prior validation study demonstrated a strong positive correlation between pulse oximetry measurement and both counting time (r = 0.59; P < 0.001) and maximal count (r = 0.67; P <0.001) achieved in one breath.7

### Outcomes

The reference measure was SpO2 as measured by a ChoiceMMed pulse oximeter (model MD300C20). Hypoxemia was defined as SpO2 of less than 95%. When SpO2 was reported over a range, the lowest value in the range was used in the analysis. Patients received instructions on correct pulse oximeter use and were told to wait 5 to 10 seconds for readings to calibrate prior to recording the SpO2 measurements.

### Statistical Analysis

In the primary analysis, the subjective dyspnea measures were treated as diagnostic tests, and the specificity (SP), sensitivity (SN), positive predictive value (PPV), negative predictive value (NPV), positive likelihood ratio (+LR) and negative likelihood ratio (-LR) were determined in order to evaluate the predictive value in detecting hypoxemia. For the continuous predictors, the test characteristics were provided across a range of different score thresholds.

Diagnostic test characteristics of the primary dyspnea measure were also determined in subgroups stratified based on patient characteristics, including (1) age <60 vs >60 years, (2) presence vs absence of underlying lung disease, and date from symptom onset (<7 vs >7 days). The Wilson method with continuity correction was used to calculate 95% confidence intervals.

A secondary analysis examined the strength of association between the presence of dyspnea and hypoxemia with a χ^2^ test or Fisher exact test with dyspnea treated as a binary variable (present or absent). This relationship is represented by a violin plot. In additional analyses, a correlation coefficient was calculated to assess whether there was an association between the participants’ Roth Scores and their oxygen saturation measurements. These associations were displayed graphically with a scatter plot. The relationship between the mMRC Dyspnea Scale and hypoxemia was analyzed with a χ^2^ test and represented by violin plot.

All analyses were conducted in SAS Statistical Software V.9.3 (Cary, North Carolina, USA).

For all statistical analyses, P < 0.05 was considered significant.

### Sample Size Calculation

The primary test characteristic of interest was the SN of dyspnea as a test for hypoxemia. The sample size was estimated based on a test of single proportion, namely SN. It was estimated that 62 patients would be required in order to estimate a SN with a 10% margin of error and 95% confidence if the true sensitivity was 80%.

## Results

### Demographic and Clinical Characteristics of Outpatients with COVID-19

From April 3 to June 8, 2020, a total of 298 patients with COVID-19 were followed by COVIDEO, 235 (78.9%) of whom were diagnosed and initially monitored in the outpatient setting. A total of 64 patients (21.5%) met criteria for home oxygen saturation monitoring. Of these patients, 59 (19.8%) were initially assessed by COVIDEO and 5 (1.7%) were followed by COVIDEO after discharge from an initial hospitalization. One patient was lost to follow up after provision of the oxygen monitoring device.

Overall, the median age of patients was 53 years (interquartile range (IQR) 38-61 years) and 41 patients (64.1%) were female (Table 1). The median number of days from symptom onset to clinical assessment was 6 (IQR 3-8). Among these patients, the most common comorbidities were hypertension (34.3%), obesity (25.0%), diabetes (17.2%), and asthma (17.2%). Nineteen patients (29.7%) had no comorbidities. The most common symptoms reported on intake assessment were cough (62.5%), fatigue/malaise (62.5%), and myalgia (45.3%). While the patients were being followed by COVIDEO, 11 (17.2%) required hospitalization, with a median duration of hospitalization of 3 days (IQR 2.5-7). Five (7.8%) patients were admitted to the intensive care unit (ICU), and the median length of ICU stay was 6 days (IQR 2-11). One patient was intubated, and no patients died within 30 days of their COVID-19 diagnosis.

**Table 1:** Demographics and Clinical Characteristics Among Outpatients with COVID-19 Monitored with Home Oxygen Saturation Devices.

|  |  |
| --- | --- |
| **Demographic information** | **No. (%)** |
| Total No. | 64 |
| Age, median (IQR), y | 53 (38-61) |
| Sex |  |
| Female | 41(64.1) |
| Male | 23 (35.9) |
| Pregnant | 3 (4.7) |
| Days from symptom onset to clinical assessment, median (IQR) | 6 (3-8) |
| **Comorbidities** |  |
| Cardiac disease | 5 (7.8) |
| Chronic lung disease | 3 (4.7) |
| Asthma | 11 (17.2) |
| Chronic kidney disease | 3 (4.7) |
| Moderate/severe liver disease | 1 (1.6) |
| Chronic neurological issues | 3 (4.7) |
| Malignancy | 9 (14.1) |
| Chronic hematological disease | 7 (10.9) |
| Diabetes | 11 (17.2) |
| Hypertension | 22 (34.3) |
| Rheumatic disorder | 2 (3.1) |
| Malnutrition | 1 (1.6) |
| Obesity | 16 (25.0) |
| None | 19 (29.7) |
| **Signs and symptoms on intake assessment** |  |
| Fever | 26 (40.6) |
| Sore Throat | 19 (29.7) |
| Runny Nose | 22 (34.4) |
| Cough | 40 (62.5) |
| Shortness of Breath | 18 (28.1) |
| Chills/Rigors | 24 (37.5) |
| Conjunctivitis | 9 (14.1) |
| Ear Pain | 6 (9.4) |
| Anosmia | 16 (25.0) |
| Dysgeusia | 19 (29.7) |
| Sputum | 9 (14.1) |
| Hemoptysis | 0 |
| Wheezing | 6 (9.4) |
| Chest Pain | 15 (23.4) |
| Myalgia | 29 (45.3) |
| Arthralgia | 11 (17.2) |
| Abdominal Pain | 10 (15.6) |
| Nausea/Vomiting | 14 (21.9) |
| Diarrhea | 18 (28.1) |
| Adenopathy | 0 |
| Rash | 1 (1.6) |
| Fatigue/Malaise | 40 (62.5) |
| Headache | 27 (42.2) |
| Confusion | 4 (6.3) |
| Depression/Anxiety | 11 (17.2) |
| Insomnia | 13 (20.3) |
| Anorexia | 24 (37.5) |
| **Laboratory findings at admission, median (IQR)** |  |
| Leukocytes, x10<sup>9</sup>/L (n=21) | 4.9 (4.5-6.1) |
| Lymphocytes, x10<sup>9</sup>/L (n=21) | 0.9 (0.7-1.2) |
| Lactate Dehydrogenase, IU/L (n=4) | 273.5 (261.3-335.5) |
| D-dimer, mcg/L (n=5) | 672.0 (516.0-796.0) |
| High-sensitivity Troponin T ng/L (n=11) | 7.5 (6.3-9.5) |
| Ferritin, mcg/L (n=3) | 986.0 (592.0-3179.0) |
| **Chest radiography done** | 21 (32.8) |
| Abnormal | 16 (76.2) |
| Bilateral infiltrates | 11 (52.4) |
| **Outcome** |  |
| ICU admission | 5 (7.8) |
| Length of ICU stay, median (IQR), days | 6 (2-11) |
| Intubation | 1 (1.6) |
| Duration of intubation, days | 15 (23.4) |
| Hospitalization | 11 (17.2) |
| Duration of hospitalization, median (IQR), days | 3 (2.5-7) |
| Multiple hospitalizations | 2 (3.1) |
| Death (within 30 days of diagnosis) | 0 |

### Association of Dyspnea Measurements with Detection of Hypoxemia

A total of 14 (22.2%) patients were diagnosed with hypoxemia. Hypoxemia was significantly associated with the presence of dyspnea (p= 0.013), mMRC Dyspnea Scale score over 0 (p= 0.02), over 1 (p= 0.0016) and over 2 (p= 0.0016) (Table 2; Figure 1). Weak associations were identified between patients’ Roth Scores and their oxygen saturation measurements for maximum count (r =0.2886; p=0.23) and counting time (r =0.1226; p=0.617), respectively.

**Table 2:**
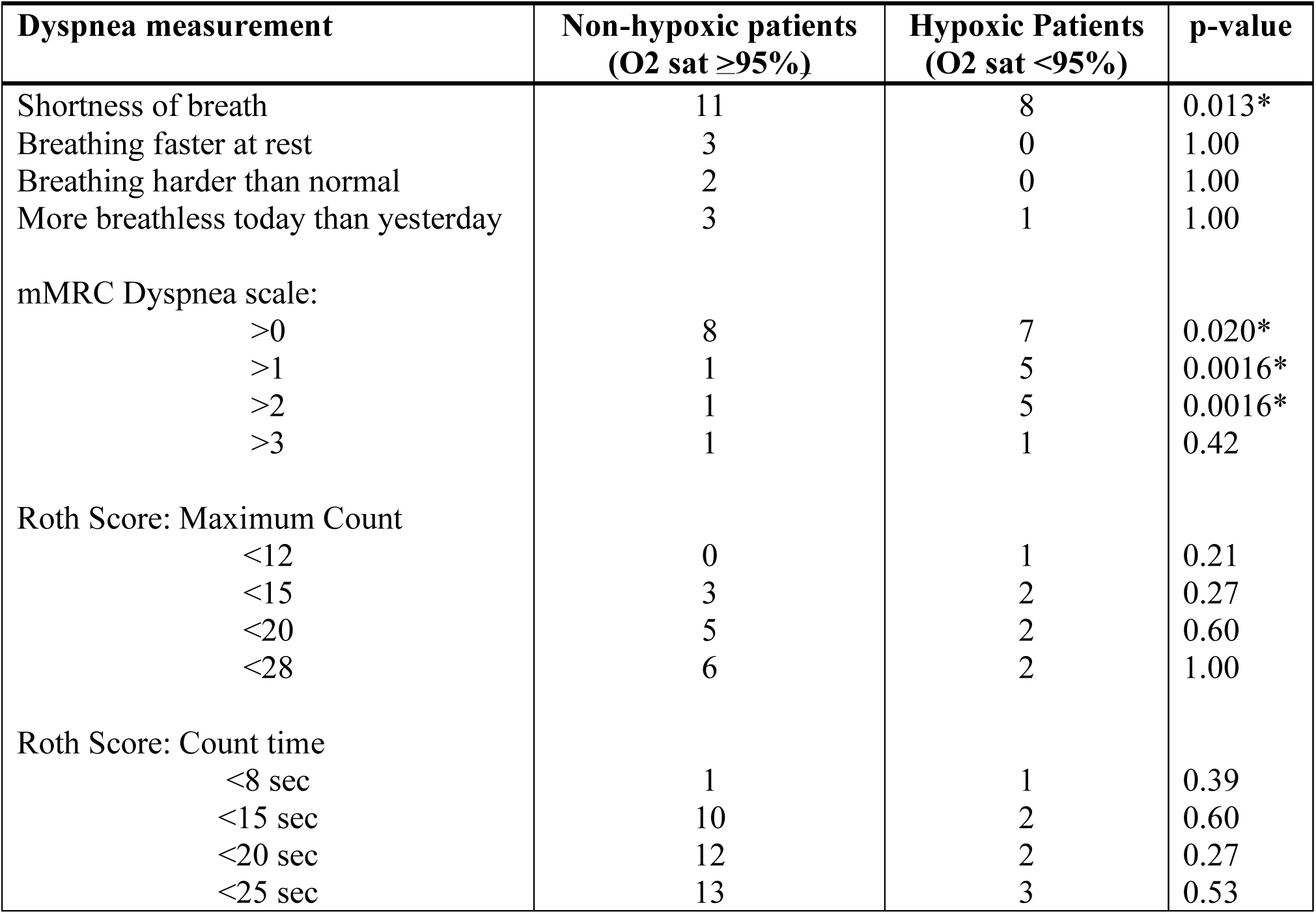
Association of Dyspnea Measurements with Detection of Hypoxemia.

|  |  |  |  |
| --- | --- | --- | --- |
| **Dyspnea measurement** | **Non-hypoxic patients**<br>**(O2 sat ≥95%)** | **Hypoxic Patients**<br>**(O2 sat <95%)** | **p-value** |
| Shortness of breath | 11 | 8 | 0.013* |
| Breathing faster at rest | 3 | 0 | 1.00 |
| Breathing harder than normal | 2 | 0 | 1.00 |
| More breathless today than yesterday | 3 | 1 | 1.00 |
| mMRC Dyspnea scale: |  |  |  |
| >0 | 8 | 7 | 0.020* |
| >1 | 1 | 5 | 0.0016* |
| >2 | 1 | 5 | 0.0016* |
| >3 | 1 | 1 | 0.42 |
| Roth Score: Maximum Count |  |  |  |
| <12 | 0 | 1 | 0.21 |
| <15 | 3 | 2 | 0.27 |
| <20 | 5 | 2 | 0.60 |
| <28 | 6 | 2 | 1.00 |
| Roth Score: Count time |  |  |  |
| <8 sec | 1 | 1 | 0.39 |
| <15 sec | 10 | 2 | 0.60 |
| <20 sec | 12 | 2 | 0.27 |
| <25 sec | 13 | 3 | 0.53 |

**Figure 1.**
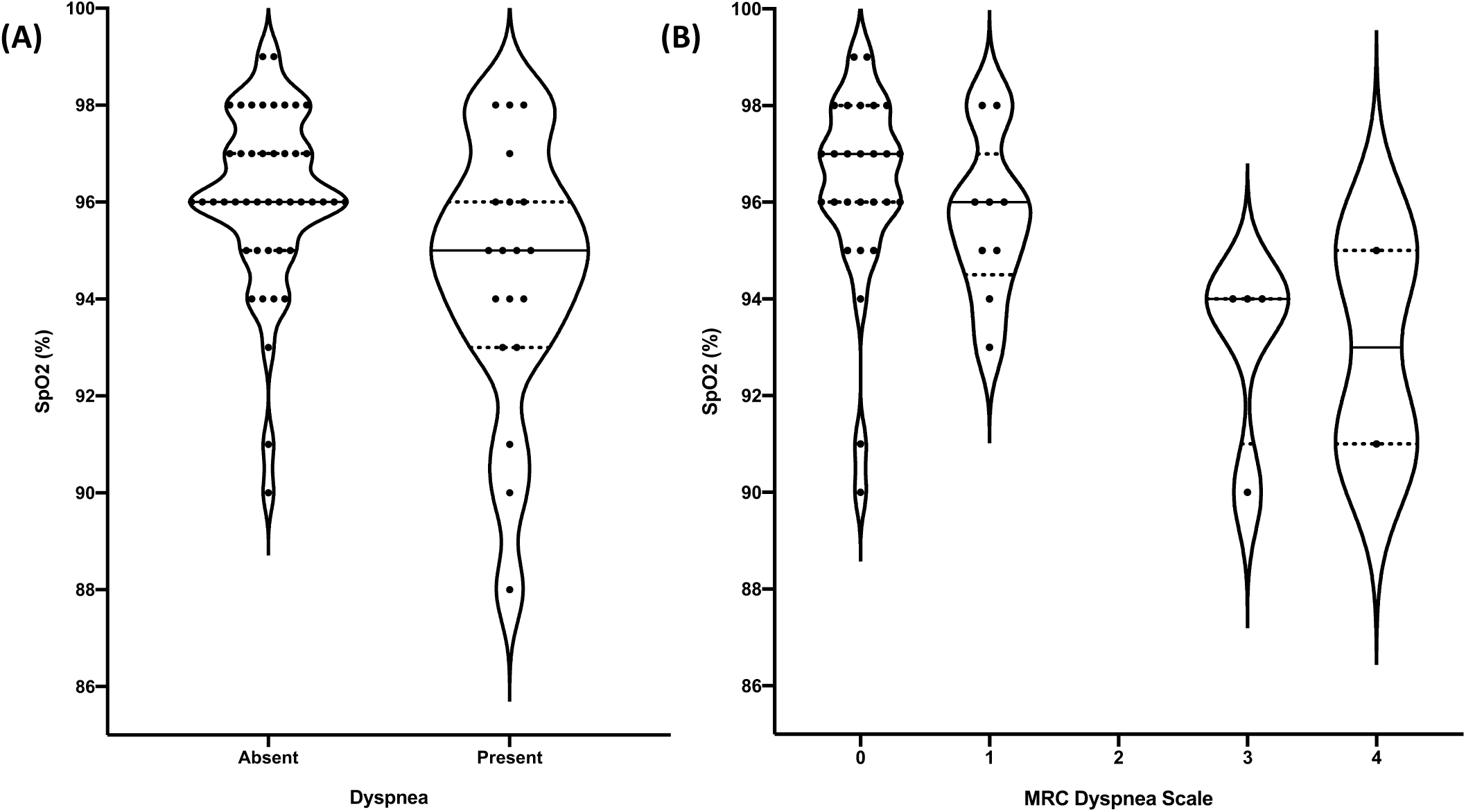
Comparison of SpO2 and measures of subjective dyspnea. A) Violin plots showing the distribution of SpO2 (%) values in COVID-19 outpatients who reported dyspnea and those who did not. B) Violin plots showing the distribution of SpO2 (%) values in COVID-19 outpatients with various mMRC Dyspnea Scale scores. The width of each plot is proportional to the number of patients with the respective SpO2 (represented by black dots). The median SpO2 is indicated by the central horizontal black line and the dotted lines correspond to the interquartile range.

### Diagnostic Accuracy of Dyspnea Measurements in the Detection of Hypoxemia

The presence or absence of subjective dyspnea had a SN 57% (95% CI 30-81%), SP 78% (63%-88%), NPV 86% (72%-94%), PPV 42% (21%-66%), +LR 2.55 (1.3-5.1), -LR 0.55 (0.3-1.0) for diagnosing hypoxemia (Table 3). The other binary measures of subjective dyspnea, including breathing faster at rest, breathing harder than normal, and feeling more breathless than the day before had lower SN (0% [0%-28%], 0% [0%-28%], and 7.7% [0.4%-38%], respectively), and higher SP (94% [82%-98%], 96% [85%-99%], and 94% [82%-98%], respectively) (Table 3).

**Table 3:** Diagnostic Accuracy of Dyspnea Measurements in the Detection of Hypoxemia: Estimation of test characteristics including sensitivity (SN), specificity, negative (NPV) and positive (PPV) predictive value and negative (-LR) and positive (+LR) likelihood ratio.

|  |  |  |  |  |  |  |
| --- | --- | --- | --- | --- | --- | --- |
| **Dyspnea measurement** | **SN**<br>**% (95%CI)** | **SP**<br>**% (95%CI)** | **NPV**<br>**% (95%CI)** | **PPV**<br>**% (95%CI)** | **-LR**<br>**(95%CI)** | **+LR**<br>**(95%CI)** |
| Shortness of breath | 57 (30-81) | 78 (63-88) | 86 (72-94) | 42 (21-66) | 0.55 (0.3-1.0) | 2.55 (1.3-5.1) |
| Breathing faster at rest | 0 (0-28) | 94 (82-98) | 78 (65-87) | 0 (0-69) | 1.07 (1.0-1.1) | 0 |
| Breathing harder than normal | 0 (0-28) | 96 (85-99) | 78 (66-88) | 0 (0-80) | 1.04 (1.0-1.1) | 0 |
| More breathless today than yesterday | 7.7 (0.4-38) | 94 (82-98) | 79 (66-88) | 25 (1.3-78) | 0.98 (0.8-1.2) | 1.26 (0.1-11.1) |
| **mMRC Dyspnea scale:** |  |  |  |  |  |  |
| >0 | 70 (35-92) | 75 (56-88) | 89 (70-97) | 47 (22-73) | 0.40 (0.2-1.1) | 2.80 (1.4-5.8) |
| >1 | 50 (24-76) | 97 (82-100) | 86 (70-95) | 83 (37-99) | 0.52 (0.3-1.0) | 16.0 (2.1-121) |
| >2 | 50 (24-76) | 97 (82-100) | 86 (70-95) | 83 (37-99) | 0.52 (28-96) | 16.0 (2.1-121) |
| >3 | 10 (0.5-46) | 97 (82-100) | 78 (61-89) | 50 (9.5-91) | 0.93 (75-115) | 3.20 (0.2-46.6) |
| **Roth Score:**<br>**Maximum Count** |  |  |  |  |  |  |
| <12 | 25 (1.3-78) | 100 (75-100) | 83 (58-96) | 100 (6-100) | 0.75 (0.4-1.3) | N/A |
| <15 | 50 (15-85) | 80 (51-95) | 86 (56-98) | 40 (7-83) | 0.63 (0.2-1.7) | 2.5 (0.6-10.2) |
| <20 | 50 (15-85) | 67 (39-87) | 83 (51-97) | 29 (5.1-70) | 0.75 (0.3-2.1) | 1.5 (0.5-5.1) |
| <28 | 7.7 (0.4-38) | 94 (82-98) | 79 (66-88) | 25 (1.3-78) | 0.98 (0.3-2.4) | 1.3 (0.4-4.0) |
| **Roth Score:**<br>**Count time** |  |  |  |  |  |  |
| <8 sec | 25 (1.3-78) | 93 (66-100) | 82 (56-95) | 50 (10-91) | 0.80 (0.5-1.4) | 3.75 (0.3-47.7) |
| <15 sec | 50 (15-85) | 33 (13-61) | 71 (30-95) | 17 (2.9-49) | 1.50 (0.5-5.1) | 0.75 (0.3-2.1) |
| <20 sec | 50 (15-85) | 20 (5.3-49) | 60 (17-93) | 14 (2.5-44) | 2.50 (0.6-10.2) | 0.63 (0.2-1.7) |
| <25 sec | 75 (22-99) | 13 (2.3-42) | 67 (13-98) | 19 (5.0-46) | 1.88 (0.2-15.8) | 0.87 (0.5-1.6) |

mMRC Dyspnea Scale scores were recorded and available for 42 patients (65.6%). An mMRC Dyspnea Scale score of greater than 0 was determined to have a SN of 70% (35%-92%), SP 75% (56%-88%), NPV 89% (70%-97%), PPV 47% (22-73%), -LR 0.40 (0.2-1.1), and +LR 2.80 (1.4-5.8) for the detection of hypoxemia. At higher cutoff values, the SN of the mMRC Dyspnea Scale was reduced to 50% (24%-76%) for scores greater than 1 and 2 and 10% (0.5%-46%) for scores greater than 3. The SP for mMRC Dyspnea Scale scores greater than 1, 2, and 3 at capturing hypoxemia was 97% (82%-100%).

Roth Scores were available for 19 patients (29.7%). The Roth Score had a higher SN at higher cutoff values for counting time. A maximum count of less than 12 had a SN of 25% (1.3%-78%), SP 100% (75%-100%), NPV 83% (58%-96%), PPV 100% (6%-100%), and a -LR of 0.75 (0.4-1.3). The diagnostic test with the highest SN for diagnosing hypoxemia was a Roth score maximum counting time of less than 25 seconds, which still had a SN of only 75% (22%-99%), and inadequate SP 13% (2.3%-42%), NPV 67% (13%-98%), PPV 19% (5.0%-46%), -LR 1.88 (0.2-16), and +LR 0.87 (0.48-1.6).

The diagnostic accuracy of dyspnea presence in the detection of hypoxemia was most impacted when stratified by the presence of underlying lung disease. In the patients with underlying lung disease, the SN and SP of the presence of dyspnea in detecting hypoxemia was 100% (20%-100%) and 75% (43%-93%), respectively. A lower SN (33% [6.0%-76%]) and high SP (100% [73%-100%]) was observed for patients over 60 years when results were stratified based on age. Stratifying based on days from symptom onset did not impact the diagnostic accuracy of dyspnea in detecting hypoxemia (Table 4).

**Table 4:** Diagnostic accuracy of the presence of dyspnea in the detection of hypoxemia stratified based on patient characteristics, including age, presence vs absence of underlying lung disease, and date from symptom onset.

|  | **SN**<br>**% (95%CI)** | **SP**<br>**% (95%CI)** | **NPV**<br>**% (95%CI)** | **PPV**<br>**% (95%CI)** | **-LR**<br>**(95%CI)** | **+LR**<br>**(95%CI)** |
| --- | --- | --- | --- | --- | --- | --- |
| **Age** |  |  |  |  |  |  |
| <60 y | 75 (36-96) | 69 (51-83) | 92 (73-99) | 35 (15-61) | 0.36 (0.1-1.2) | 2.39 (1.3-4.5) |
| ≥60 y | 33 (6.0-76) | 100 (73-100) | 78 (52-93) | 100 (20-100) | 0.67 (0.4-1.2) | N/A |
| **Underlying lung disease** |  |  |  |  |  |  |
| Yes | 100 (20-100) | 75 (43-93) | 100 (63-100) | 40 (7.3-83) | 0 | 4.00 (1.5-10.7) |
| No | 50 (25-75) | 78 (61-90) | 83 (66-93) | 43 (19-70) | 0.64 (0.4-1.2) | 2.31 (1.0-5.3) |
| **Days from symptom onset** |  |  |  |  |  |  |
| <7 days | 60 (17-93) | 83 (64-94) | 92 (73-99) | 38 (10-74) | 0.48 (0.2-1.4) | 3.48 (1.2-10.2) |
| ≥7 days | 63 (26-90) | 71 (44-89) | 80 (51-95) | 50 (24-76) | 0.53 (0.2-1.4) | 2.1 (0.9-5.3) |

## Discussion

To our knowledge, this is the first study to evaluate the diagnostic accuracy of subjective dyspnea in detecting hypoxemia in the setting of COVID-19. Self-reported shortness of breath has very limited utility for detecting hypoxemia, with a SN of only 57% and SP of only 78% for detecting O2 saturation levels below 95%. An mMRC Dyspnea Score exceeding 1, a Roth maximal count less than 12, and Roth counting time under 8 seconds offered high SP and +LR to rule in hypoxemia. Identifying patients with these features may be helpful in the remote assessment of COVID-19 outpatients. However, none of these measures offered sufficient SN or –LR to help rule out hypoxemia–which is the more clinically important consideration for these patients.

Previous studies examining the correlation between subjective dyspnea and hypoxemia in other respiratory conditions have yielded inconsistent findings. The strongest confirmation of the potential diagnostic utility of dyspnea emerged from a study of 76 patients admitted to the emergency department with acute exacerbations of COPD, in which dyspnea scores exceeding 3 or 4 on a 5-stage scoring system were found to have a sensitivity of 93.5% for detecting hypoxemia.^13^ Additionally, the mMRC Dyspnea Scale has been found to be significantly correlated with SpO2 in measurements obtained during exercise among patients with idiopathic pulmonary fibrosis.^18^ Conversely, several other studies have shown no correlation between perceived dyspnea and hypoxemia in conditions such as advanced lung cancer, COPD, and palliative care patients.^14-15,19^

Discrepancies between respiratory rate and SpO2 in COVID-19 patients with acute respiratory failure have been highlighted previously, suggesting that a normal respiratory rate may belie profound hypoxemia in this setting.^20^ High levels of anxiety may contribute to feelings of dyspnea in patients who are non-hypoxemic. There are also a growing number of case reports documenting silent hypoxemia among COVID-19 patients, where patients present with hypoxemia in the absence of respiratory symptoms.^9^-^10^, ^21^ The underlying mechanism responsible for severe hypoxemia in the absence of dyspnea is not well elucidated. It has been postulated that this clinical picture may be consistent with a phenotype of COVID-19 pneumonia (L-phenotype) characterized by low elastance, low ventilation-perfusion ratio and near normal compliance.^11^ The relatively high compliance results in preserved gas volumes, while hypoxemia may result due to a ventilation-perfusion mismatch caused by impaired lung perfusion regulation and loss of hypoxic vasoconstriction. ^22-23^ Additionally, the absence of dyspnea despite severe hypoxemia may reflect pulmonary vaso-occlusive disease, whereby patients develop clinically silent microvascular thrombi in early stages of the disease, which if left untreated, results in worsening hypercoagulability and rapid clinical deterioration due to a thrombo-inflammatory cascade.^24-25^ While at this point the exact mechanism remains speculative, our data suggest that the discrepancy between dyspnea and hypoxemia makes it difficult to accurately assess patients remotely and emphasizes the importance of SpO2 monitoring in order to avoid missing patients with developing respiratory failure.

This study has several limitations. The data collected for this study were from patients’ initial pulse oximeter assessment and did not assess whether changes in dyspnea correlate with changes in SpO2 over time. This is a potentially important notion when monitoring patients who are (or are not) becoming increasingly dyspneic while self-isolating in their homes. While the number of patients included was sufficient for the primary analysis, they were insufficient for precise estimates of subgroups stratified by age, presence of lung disease, and date of symptom onset. Additionally, our study was limited to patients who were considered at high risk of severe disease, and it is possible that the diagnostic test characteristics might differ in younger and healthier patients. Lastly, pulse oximeters may have variable accuracy as individuals become increasingly hypoxic and are further impacted by individual patient characteristics; however, a perfect reference standard of invasive blood oxygen measurement would be neither practical nor ethical in the outpatient setting^.26^

Our findings indicate that subjective dyspnea does not accurately capture hypoxemia in patients with COVID-19. Although some dyspnea scores have high specificity and positive likelihood ratios for identifying hypoxemia, none of these measures have sufficient sensitivity to rule out hypoxemia. Therefore, relying on surrogate measures of dyspnea alone is not sufficient to remotely monitor high-risk outpatients with COVID-19. Home SpO2 monitoring should be a mandatory component of remote management all high-risk outpatients with COVID-19.

## Data Availability

N/A

## Author Contributions

The authors all stand behind the conclusions of this manuscript, agree to be accountable for all aspects of the work, and support its publication. All authors contributed to the study conception and designed the protocol. All authors contributed to the manuscript preparation and have given approval for its submission.

## Conflicts of Interest

The authors report no financial or other conflicts of interest.

## Supplementary Material

**Figure S1.**
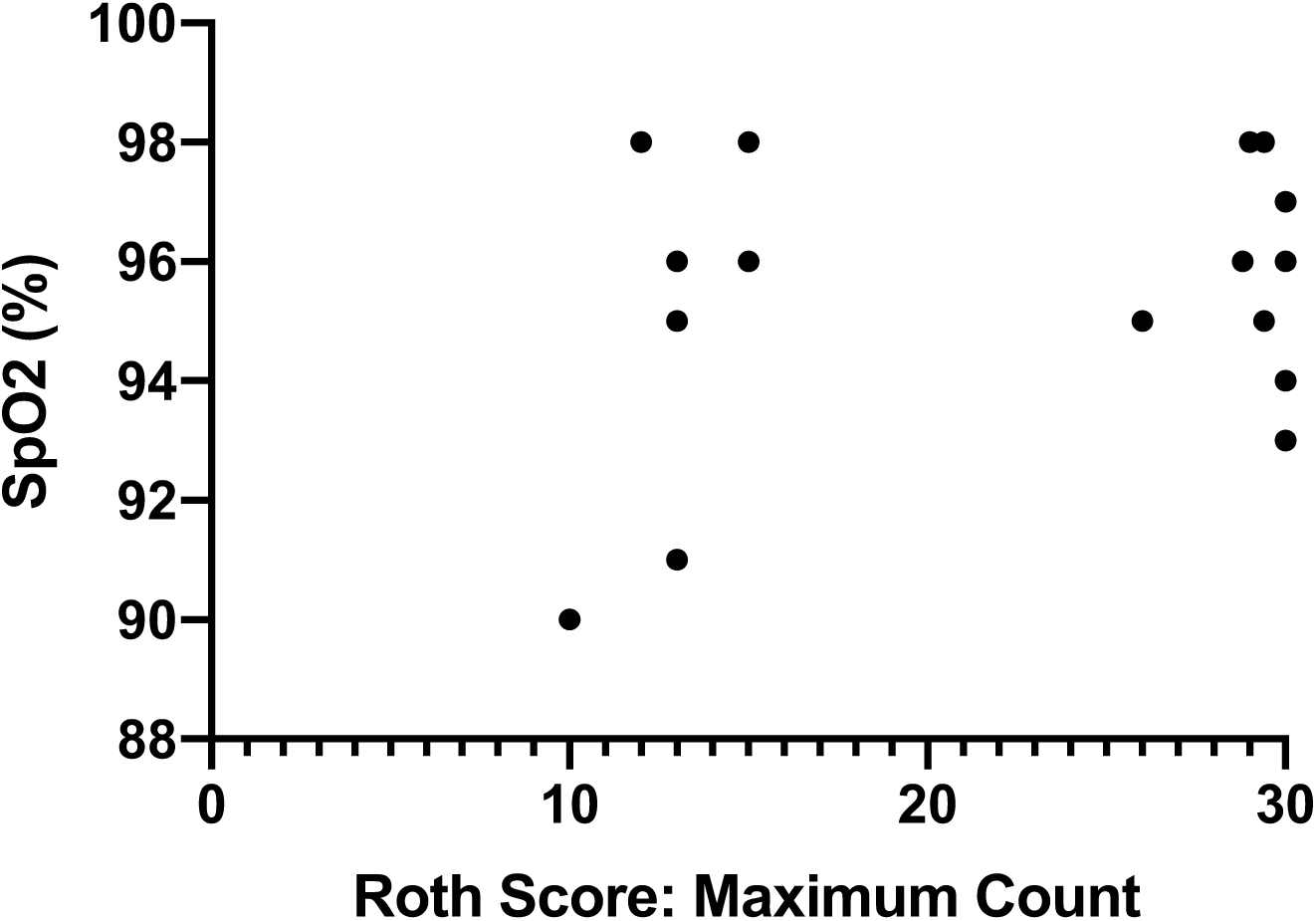
Scatter Plot of SpO2 values across patients’ Roth Scores (Maximum Count)

**Figure S2.**
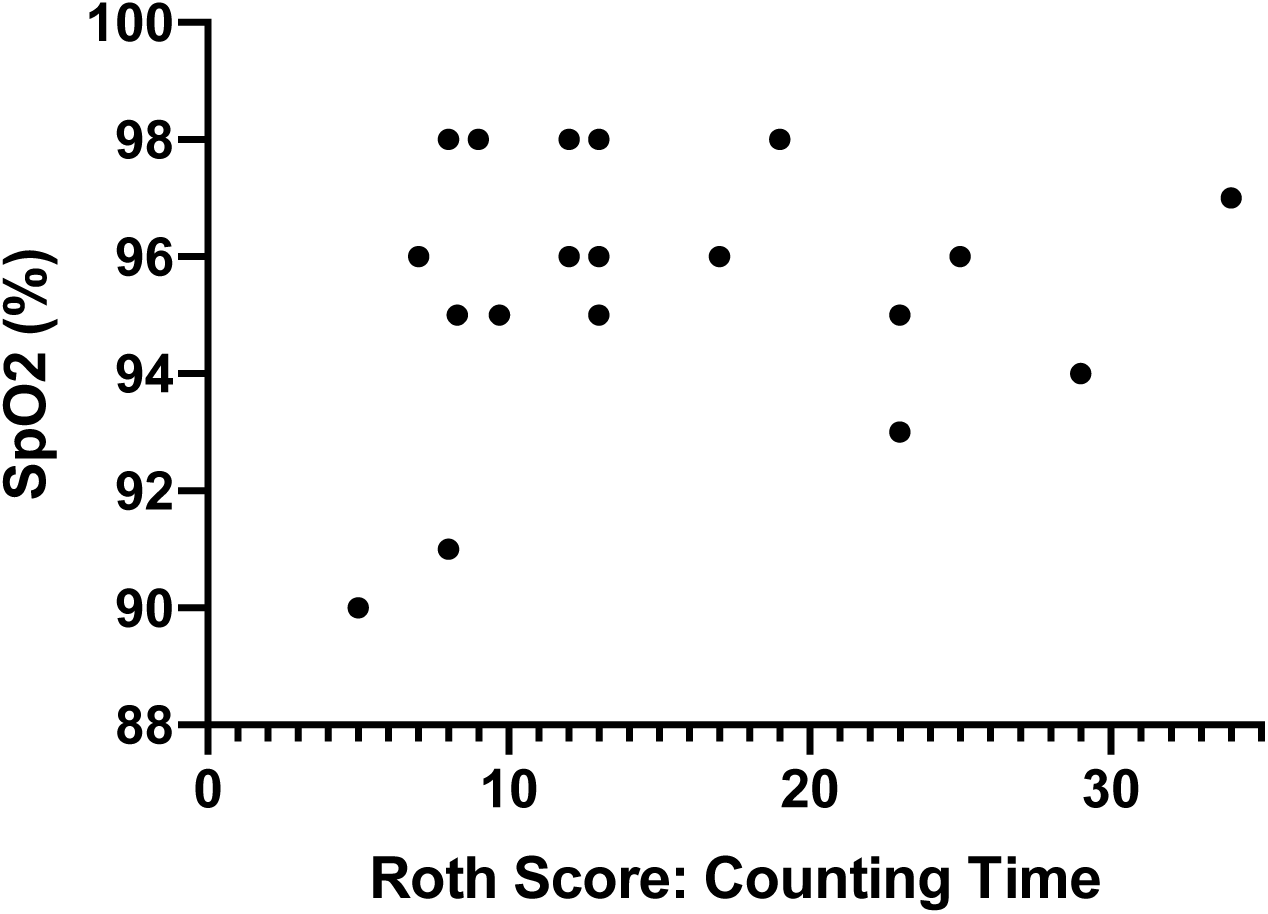
Scatter Plot of SpO2 values across patients’ Roth Scores (Counting Time)

## Notes

### Competing Interest Statement

The authors have declared no competing interest.

### Author Declarations

This study was approved by the institutional review board at Sunnybrook Health Sciences as minimal risk research, using data collected for routine clinical practice, and the requirement for informed consent was waived

